# Spatial Clustering of School Susceptibles Drives Divergent US Measles Outbreaks

**DOI:** 10.64898/2026.02.25.26347103

**Authors:** Siyu Chen, Nathaniel Hupert, Ana I. Bento

## Abstract

The two largest US measles outbreaks in over two decades (2025 Gaines County, Texas: 414 cases, contained; 2025-2026 Spartanburg County, South Carolina: 923+ cases, ongoing) occurred in counties with similar sub-threshold K-12 MMR coverage (85.1% vs 88.8%), yet their trajectories diverged dramatically. Using kernel density estimation with a common bandwidth and bootstrap uncertainty quantification, we compared sub-county vaccination data at the district level for Texas (3 districts, 3,560 students) and the school level for South Carolina (93 schools, 57,281 students). Peak susceptible density in Spartanburg County was 5.7 times that of Gaines County (23 vs. 4 unvaccinated students per square mile; 95% CI 2.4-12.5, non-overlapping). In Texas, a single isolated cluster around Seminole ISD limited spatial connectivity, producing self-limiting spread. In South Carolina, a northwest corridor of under-vaccinated schools created overlapping catchment areas that sustained transmission chains. These findings demonstrate how county-level aggregates can mask nearly six-fold differences in local risk, underscoring the need for school-level spatial surveillance.

## Introduction

Measles outbreaks persist not because of average vaccination coverage shortfalls but because of the local spatial density of susceptible individuals [1]. When unvaccinated children cluster geographically, effective reproduction numbers can exceed the epidemic threshold even in communities with nominally adequate coverage [2, 3]. For measles, with a basic reproduction number of 12-18, the critical vaccination threshold exceeds 95%, making transmission dynamics acutely sensitive to spatial heterogeneity in the distribution of unvaccinated individuals [4]. As US county-level Measles-Mumps-Rubella (MMR) coverage has declined [5] and exemption rates have risen [6], analyses relying on county-scale data miss the sub-county clustering of susceptibles that theory predicts should drive outbreak growth and persistence [3,7]. Simulation studies confirm that fine-scale clustering amplifies outbreak potential in ways obscured by aggregates, yet granular vaccination data in the US have been too sparse to validate these predictions empirically [3].

Scale matters: county-level surveillance obscures the very clustering that drives transmission dynamics. The 2025 Gaines County, Texas outbreak (414 cases, contained) and the 2025–2026 Spartanburg County, South Carolina outbreak (923+ cases, ongoing) representing the two largest US measles outbreaks in over two decades, had similar sub-threshold K-12 MMR coverage (85.1% vs. 88.8%). We hypothesized that the sub-county spatial density of susceptible students, not county-level coverage alone, explains their divergent trajectories. Using sub-county vaccination data and kernel density estimation (KDE), we quantified a 5.7-fold difference in peak susceptible density (23 vs. 4 unvaccinated students per square mile) that county aggregates entirely mask.

## Results

K-12 MMR vaccination coverage averaged 85.1% in Gaines County, Texas and 88.8% in Spartanburg County, South Carolina (Figure 1A). Coverage varied by grade level in both counties, with elementary schools showing the lowest rates (81.0% and 86.8%, respectively) and high schools the highest (90.2% and 93.1%). Although Spartanburg County’s overall coverage exceeded that of Gaines County by 3.7 percentage points, these county-level averages masked dramatically different sub-county susceptibility landscapes.

**Figure 1.**
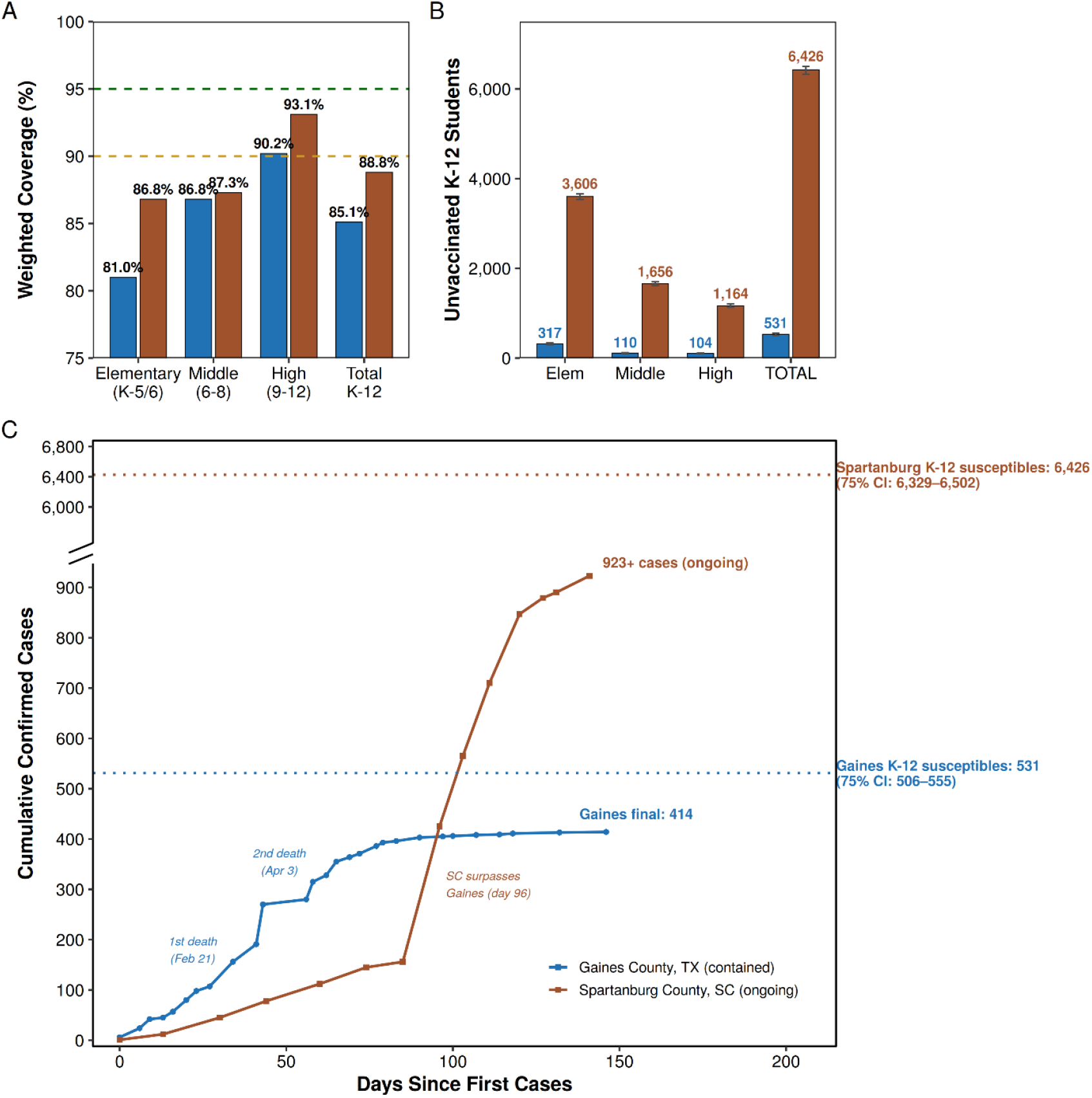
Sub-county vaccination coverage, susceptibility, and outbreak trajectories in Gaines County, Texas (contained outbreak) and Spartanburg County, South Carolina (ongoing outbreak) (A) Weighted MMR vaccination coverage by school level for Gaines County (blue) and Spartanburg County (brown): elementary (K-5/6), middle (6-8), high (9-12), and total K–12. Both counties fall below the 95% herd immunity threshold (green dashed line) and the 90% surveillance target (orange dashed line) at the total K-12 level, with elementary coverage lowest in both (81.0% vs 86.8%). (B) Estimated number of unvaccinated K-12 students by school level in each county, with 75% bootstrap confidence intervals. Spartanburg has 12-fold more unvaccinated students overall (6,426 vs 531), largely reflecting its 16-fold larger enrollment. (C) Cumulative confirmed measles cases over days since first reported case. Horizontal dashed lines indicate estimated K-12 susceptible population ceilings for each county with 75% CIs (Gaines: 531, 506-555; Spartanburg: 6,426, 6,329-6,502). The Gaines outbreak plateaued at 414 reported cases after approximately 147 days; the Spartanburg outbreak surpassed Gaines at day 96 and continues with 923+ reported cases after 200+ days.

Within Gaines County, sub-county variation was dramatic: coverage ranged from 60% to 96% across eight schools in three districts, with Loop ISD at the lowest end (Figure 2A). Spartanburg County showed a wider spread: 21% to 100% across 93 schools, with 24 schools (26%) falling below 90% (Figure 2B). We estimated 531 (75% CI: 506-555) unvaccinated K-12 students in Gaines County and 6,426 (75% CI: 6,329-6,502) in Spartanburg County (Figure 1B), a 12-fold gap largely reflecting the 16-fold difference in enrollment (3,560 vs 57,281).

**Figure 2.**
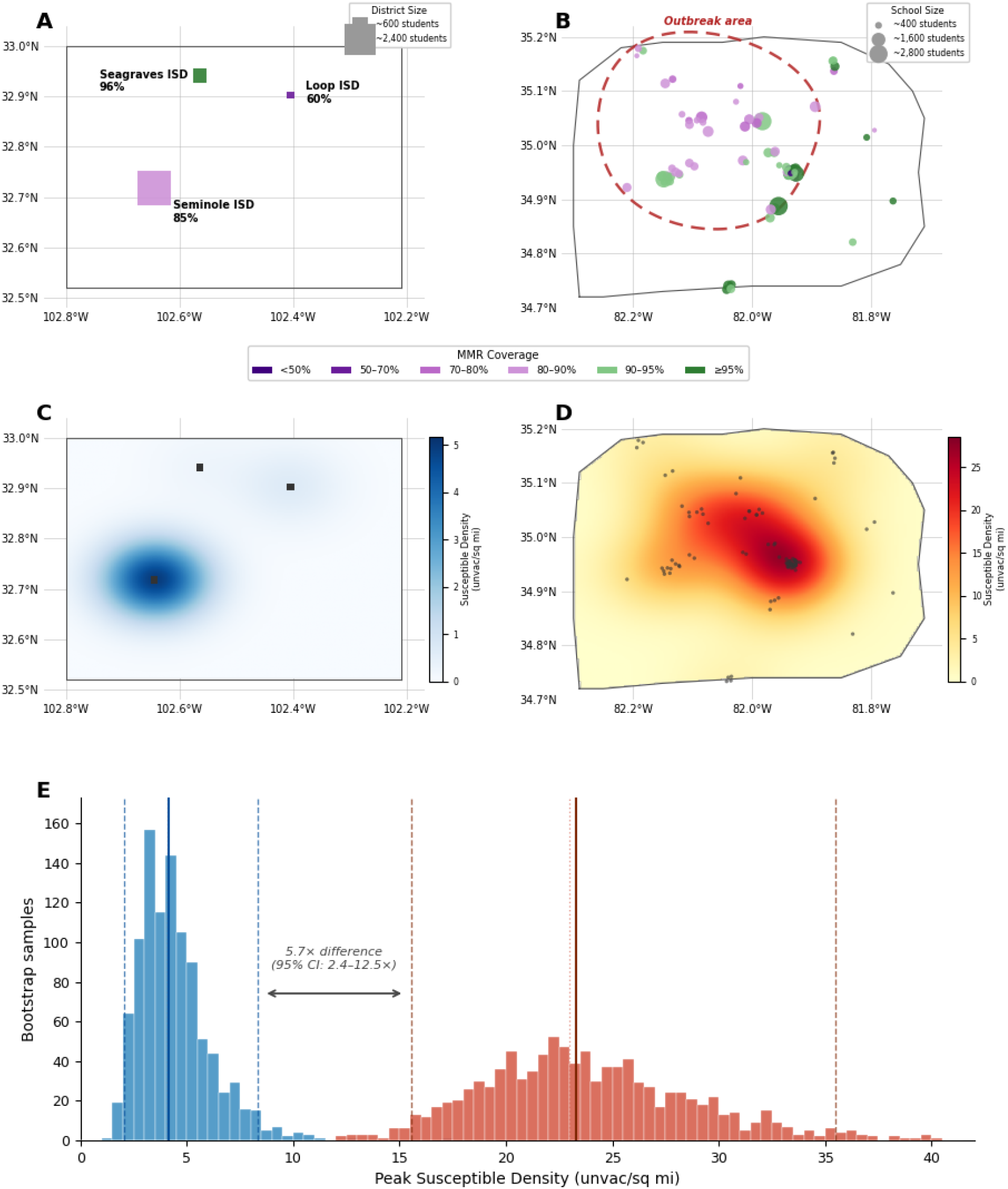
Spatial heterogeneity in sub-county vaccination coverage and susceptible population density among K–12 students. (A) Gaines County, Texas: district-level MMR coverage for three independent school districts; symbol size proportional to enrollment (∼600 to ∼2,400 students) and color indicates coverage category (<50% to ≥95%). Loop ISD (60% coverage, 149 students) is geographically isolated in the northeast. (B) Spartanburg County, South Carolina: school-level MMR coverage for 93 schools; symbol size proportional to enrollment (∼400 to ∼2,800 students). Under-vaccinated schools cluster in the northwestern quadrant; the dashed red boundary delineates the convex hull of 33 confirmed exposure school sites [12]. (C) Kernel density estimation (KDE) of susceptible K-12 students in Gaines County, showing unvaccinated students per square mile using a common 7-km Gaussian bandwidth. Peak density concentrated around Seminole ISD with maximum ∼5 unvaccinated per square mile. (D) KDE of susceptible K-12 students in Spartanburg County using the same 7-km bandwidth. Peak density in the northwest cluster exceeds 20 unvaccinated per square mile. Note different color scales between (C) and (D). (E) Bootstrap distribution (n = 1,000) of peak susceptible density for Gaines County (blue; median 4.1 unvaccinated/sq mi; 95% CI: 2.1-8.3) and Spartanburg County (red; median 23 unvaccinated/sq mi; 95% CI: 16-35). Solid vertical lines indicate medians; dashed lines indicate 2.5th and 97.5th percentiles. The median ratio of peak densities is 5.7× (95% CI: 2.4-12.5×).

To test whether spatial structure explains outbreak dynamics beyond total susceptible counts, we applied kernel density estimation using a common Gaussian bandwidth (0.066 degrees, approximately 7 km) to both counties. Peak susceptible density reached 23 unvaccinated students per square mile in Spartanburg County (95% CI: 16-35), versus 4.1 in Gaines County (95% CI: 2.1-8.3), a 5.7-fold difference (95% CI: 2.4-12.5) with non-overlapping confidence intervals (Figure 2C-E). The spatial distribution of susceptibles differed qualitatively. Gaines County’s single highly susceptible district (Seminole ISD) was geographically isolated from the county’s two other districts, limiting spatial susceptible connectivity. Spartanburg County’s multiple under-vaccinated schools clustered in the northwest quadrant, forming overlapping catchment areas primed for sustained transmission.

These patterns match outbreak trajectories (Figure 1C). In Gaines County, case counts plateaued after 147 days at 414 cases, corresponding to a school-age attack rate of approximately 52-78% after adjusting for the estimated one-third of cases occurring in adults. In Spartanburg County, the larger susceptible pool (∼6,426 K-12 students) is distributed across 93 schools, enabling sequential propagation through the school network. Exposure-site data show the outbreak expanding from two initial school sites in October 2025 to 33 confirmed exposure sites by January 2026, spreading progressively outward from the northwestern high-density cluster identified by KDE (dashed boundary, Figure 2B; Supplementary Extended Dataset). After 200+ days and 923+ confirmed cases, with new school sites still being identified, the Spartanburg outbreak had not yet exhausted its spatial reservoir of susceptible students, consistent with the 5.7-fold higher peak susceptible density estimated in Figure 2E.

## Discussion

Sub-county vaccination data reveal spatial heterogeneity in susceptibility that determines outbreak dynamics but remains invisible at the county level. Spartanburg County’s larger absolute susceptible pool partly reflects its bigger population, yet it is the geographic clustering of under-vaccinated schools, not merely the total number of susceptibles, that distinguishes Texas’s self-limiting outbreak from South Carolina’s sustained transmission. In Gaines County, susceptibles were concentrated in a single isolated district (Seminole ISD), forming a natural spatial barrier to onward spread. In Spartanburg County, a dense cluster of under-vaccinated schools in the northwest created overlapping catchment areas that enabled persistent transmission chains.

Our findings are consistent with earlier theoretical work [2,3]. Park et al. [2] showed that susceptible dynamics govern pathogen persistence, with measles uniquely vulnerable due to slow susceptible pool replenishment. Masters et al. [3] demonstrated through simulation that fine-scale nonvaccinating clustering generates outbreak risks hidden from aggregate surveillance, where county-level data substantially underestimate expected outbreak size. Their proof of concept was limited, however, by the absence of granular US vaccination data. We advance this one step further with a data-driven empirical test: school-level analysis of the two largest US measles outbreaks in over two decades quantifies a 5.7-fold peak density gap (23 vs. 4 unvaccinated students per square mile) that county-level metrics entirely mask, validating their theoretical predictions with observed outbreak data.

Several limitations warrant consideration. Vaccination data differ in granularity between the two counties: district-level in Texas (3 units) versus school-level in South Carolina (93 units). We frame this asymmetry as illustrating the value of finer-resolution data rather than as a limitation of comparability, and non-overlapping confidence intervals confirm that the density gap is robust to this difference; finer school-level data from Texas, if available, would likely reinforce the spatial isolation pattern. Cohort reconstruction in Texas may overestimate susceptibility if catch-up vaccination has occurred, though sensitivity analyses narrow the density gap without eliminating it. Other factors including differences in urbanization, public health response, population mixing, and case importation likely contributed to the divergent trajectories, but spatial density of susceptibles offers a parsimonious, quantifiable mechanism that county-level data alone cannot capture.

County-only reporting risks providing false reassurance where sub-county pockets of susceptibility exist. We recommend that public health agencies prioritize collection of school-level vaccination data and integrate spatial clustering analysis into routine outbreak risk assessment [8].

## Materials and Methods

We obtained sub-county MMR vaccination coverage from the Texas Department of State Health Services (2019-2025 kindergarten and 7th-grade data for 3 independent school districts [9] and the South Carolina Department of Health (2025-2026 immunization report for 93 schools, 57,281 students) [10]. For Texas, we reconstructed grade-specific susceptible cohorts using historical coverage and enrollment data during 2024-2025 school year. For South Carolina, we calculated unvaccinated students directly from school-wide immunization rates across grades and reconstructed grade-specific susceptible cohorts during 2025-2026 school year (SI Extended Dataset and Extended Methods). Different data structures require different analytical assumptions. The comparison highlights sub-county granularity rather than equivalent school-level data.

We applied Gaussian kernel density estimation weighted by unvaccinated student counts using a common bandwidth of 0.066 degrees (approximately 7 km) for both counties, converting the resulting density surfaces to unvaccinated students per square mile. We quantified uncertainty through 1,000 bootstrap iterations incorporating coverage perturbation (SD: ±3 percentage points for school-level South Carolina data, ±5 for district-level Texas data), spatial jitter, and resampling (SI Extended Methods).

Case counts and spatial information were obtained from the Johns Hopkins Measles Tracking Dashboard [11] for Gaines County and from South Carolina Department of Health reports for Spartanburg County [12]. All data were publicly available and de-identified. Data were analyzed with Python and R.

## Data Availability

All data, code, and materials are available at: https://github.com/SiyuChenOxf/USmeasles_outbreak

## Author Contributions

Chen and Bento had full access to all of the data in the study and took responsibility for the integrity of the data and the accuracy of the data analysis.

Concept and design: Chen & Bento.

Acquisition, analysis, or interpretation of data: Chen & Bento.

Drafting of the manuscript: Chen, Hupert and Bento.

Critical review of the manuscript for important intellectual content: Chen, Hupert and Bento.

Statistical analysis: Chen.

Obtained funding: Bento.

Administrative, technical, or material support: Chen & Bento. Supervision: Bento.

Data sharing: All data, code, and materials are available at: https://github.com/SiyuChenOxf/USmeasles_outbreak

## Supporting Methods

### Data Sources

#### Texas Data

School-level vaccination coverage data for Texas were obtained from the Texas Department of State Health Services (DSHS) Annual School / Facility Vaccination Coverage Level Reports for kindergarten students (2019-2020 through 2024-2025) and 7th grade students (2023-2024 through 2024-2025) [1]. These reports provide district-level and school-level measles, mumps, rubella (MMR) vaccination rates collected during the fall enrollment period each academic year.

School enrollment data by grade level were obtained from the Texas Education Agency (TEA) Statewide Campus-Level Enrollment Reports (2019-2020 through 2024-2025). These data provide student counts by grade (kindergarten through 12th grade) for each school campus [2].

List of schools within each school district in Gaines County by ISD:

### Seminole ISD (6 campuses) [3]

F.J. Young — PK–1

Seminole Primary — Grades 2–3

Seminole Elementary — Grades 4–5

Seminole Junior High — Grades 6–8

Seminole Success Center — alternative campus

Seminole High School — Grades 9–12

### Seagraves ISD (1 campus) [4]

Seagraves Schools — PK–12 (single combined campus serving all grades)

### Loop ISD (1 campus) [5]

Loop School — PK–12 (single combined campus serving all grades)

Outbreak case counts were obtained from the Johns Hopkins Measles Tracking Team dashboard [6], which aggregates data from state health department reports.

#### South Carolina Data

School-level immunization rates for South Carolina were obtained from the South Carolina Department of Health and Environmental Control (DHEC) 45-Day Immunization Report (2025-2026 school year). This report provides school-wide immunization compliance rates across all enrolled grades, not kindergarten-specific rates [7]. The dataset includes school name, county, grade range served, total enrollment, and overall immunization compliance percentage.

Outbreak case counts were obtained from the South Carolina Department of Public Health Measles Dashboard [8].

School-level exposure site data were compiled from the South Carolina Department of Public Health situation reports issued between October 2025 and February 2026. A total of 33 school sites were identified as exposure locations over the course of the Spartanburg County outbreak, comprising 17 elementary schools, 6 middle schools, 5 high schools, 2 charter K-12 schools, 1 K-8 school, and 2 universities. The earliest exposure sites were reported on October 8, 2025 (Global Academy of South Carolina and Fairforest Elementary), coinciding with outbreak day 13. Exposure site identification accelerated markedly through December 2025 and January 2026, with 12 new sites reported in December and 16 in January [8]. The peak quarantine burden was observed at Global Academy of South Carolina, where 139 students were placed under quarantine. Two university campuses (Clemson University and Anderson University) were identified as exposure sites on January 20, 2026, reflecting geographic spread beyond the K-12 school system. For each school, we recorded the date of first reported exposure, the cumulative outbreak case count at the time of first exposure, the maximum quarantine count observed, the date and case count of the most recent situation report mentioning the school, and the total number of report dates on which the school appeared. These data are provided in supplementary dataset (SC_Spartanburg_Detailed_Calculations.csv) on Github. The geographic coordinates of all 33 exposure sites were used to delineate the outbreak boundary shown in Figure 2B.

## Data Comparability

A critical methodological challenge in this analysis is that Texas and South Carolina report vaccination data differently:

- Texas: district-level reporting of kindergarten-specific and 7th-grade-specific MMR coverage rates, with historical data available from 2019 to 2025.
- South Carolina: school-level reporting of school-wide immunization rates across all grades, for the current year only (2025–2026).

To address these differences, we employed two complementary approaches: (1) stratified analysis of South Carolina schools by grade range to identify comparable age groups, and (2) cohort reconstruction methodology for Texas to estimate current grade-specific susceptibility.

### School Categorization (South Carolina)

Spartanburg County schools were categorized based on reported grade ranges into six categories:

| Category | Definition | Schools (n) | Students |
| --- | --- | --- | --- |
| Elementary (K-5/6) | Grades K through 5 or 6 | 44 | 22,619 |
| Middle (6-8) | Grades 6 through 8 only | 16 | 10,464 |
| High (9-12) | Grades 9 through 12 only | 11 | 14,930 |
| K-8 / K-6 | Full K-8 or K-6 span | 13 | 4,454 |
| K-12 | Full K-12 span | 7 | 3,247 |
| Middle-High (7-12) | Grades 7 through 12 | 2 | 1,566 |
| **TOTAL** |  | **93** | **57,280** |

Of the 93 schools in Spartanburg County, 71 are pure single-level schools (elementary, middle, or high only) and 22 are multi-grade schools spanning multiple levels (K-8, K-12, or middle-high). For the primary analysis, we report three South Carolina subsets: (1) all kindergarten-serving schools (K-including; n=64, combining elementary, K-8, and K-12), (2) elementary-only schools (K-5/6; n=44, most age-comparable to Texas kindergarten data), and (3) all schools (n=93). For multi-grade schools, enrollment and unvaccinated students were allocated proportionally across levels by grade span (e.g., a K-8 school with 9 grade levels assigns 6/9 of enrollment to elementary and 3/9 to middle). Freshman academies (Byrnes 1-9, Dorman 1-9) were classified as High (grade 9 only).

### Cohort Reconstruction Methodology (Texas)

To estimate current vaccination coverage by grade level in Gaines County, Texas, we developed a cohort reconstruction approach that applies historical kindergarten coverage rates to current enrollment:

The method assumes that vaccination status established at kindergarten entry remains relatively stable through elementary school. Current students in grades K–5 are assigned the kindergarten coverage rate from the year they entered kindergarten (e.g., current 3rd graders in 2024-25 are assigned the 2021-22 kindergarten coverage rate). Students in grades 6–12 are assigned the 7th grade coverage rate from the appropriate cohort year, as 7th grade represents the next mandated vaccination checkpoint in Texas.

#### Grade-to-Coverage Year Mapping

| Current Grade (2024-25) | Cohort Year | Coverage Source |
| --- | --- | --- |
| Kindergarten | 2024-25 | 2024-25 Kindergarten rate |
| 1st Grade | 2023-24 | 2023-24 Kindergarten rate |
| 2nd Grade | 2022-23 | 2022-23 Kindergarten rate |
| 3rd Grade | 2021-22 | 2021-22 Kindergarten rate |
| 4th Grade | 2020-21 | 2020-21 Kindergarten rate |
| 5th Grade | 2019-20 | 2019-20 Kindergarten rate |
| 6th Grade | 2024-25 | 2024-25 7th grade rate |
| 7th Grade | 2024-25 | 2024-25 7th grade rate |
| 8th Grade | 2023-24 | 2023-24 7th grade rate |
| 9th Grade | 2022-23 | 2022-23 7th grade rate |
| 10th Grade | 2021-22 | 2021-22 7th grade rate |
| 11th Grade | 2020-21 | 2020-21 7th grade rate |
| 12th Grade | 2019-20 | 2019-20 7th grade rate |

### Susceptible Density Estimation

To quantify and compare the geographic distribution of unvaccinated students between outbreak regions, we used kernel density estimation (KDE) to create continuous density surfaces from discrete school locations.

#### Estimating Unvaccinated Students by School

For Texas, we calculated unvaccinated students at each school district using cohort-reconstructed coverage rates:

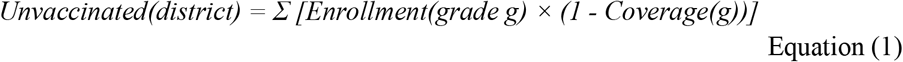

where coverage for each grade was assigned based on the cohort mapping table above (K rates for grades K–5, 7th grade rates for grades 6–12).

For South Carolina, we calculated unvaccinated students directly from school-wide data:

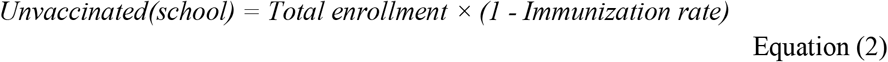

#### Uncertainty in Susceptible Estimates

To quantify uncertainty in the estimated number of unvaccinated students, we used a parametric bootstrap procedure (n = 1,000 iterations). In each iteration, the coverage rate for each reporting unit (district for Texas, school for South Carolina) was perturbed by drawing from a normal distribution centered on the observed rate, with standard deviations of ±5 percentage points for Texas district-level data (reflecting additional uncertainty from cohort reconstruction) and ±3 percentage points for South Carolina school-level data (reflecting measurement error in school-wide immunization rates). Perturbed coverage rates were truncated to [0, 1]. Given the perturbed coverage rate, the number of unvaccinated students at each unit was resampled from a binomial distribution:

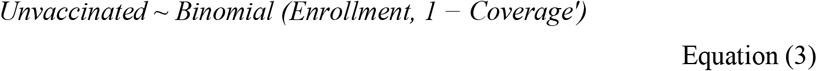

where Coverage′ is the perturbed rate. County-level totals were computed by summing across all units within each county, and level-specific totals (elementary, middle, high) were computed by summing within each school level. We report 75% confidence intervals (12.5th and 87.5th percentiles) for the susceptible estimates shown in Figure 1B-C, yielding: Gaines County K-12 total 531 (75% CI: 506–555) and Spartanburg County K-12 total 6,426 (75% CI: 6,329-6,502). The narrower relative width of the Spartanburg interval reflects the larger number of reporting units (93 schools vs 3 districts), which averages out unit-level perturbations.

#### Kernel Density Estimation

School locations were geocoded using school addresses. We then applied Gaussian kernel density estimation to create continuous density surfaces, with each school location weighted by its estimated number of unvaccinated students.

The kernel density estimator calculates a smooth density function *f* (*x*) across geographic space:

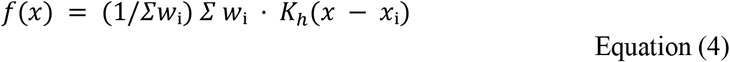

where *K* is the Gaussian kernel function, *h* is the bandwidth, *x*_i_ are school coordinates, *w*_i_ is the weight (number of unvaccinated students at school *i*), and *n* is the total number of schools.

The resulting density surfaces were normalized to represent unvaccinated students per square mile, enabling direct comparison between counties despite differences in total area and student population.

KDE Bandwidth Selection. Gaussian kernel bandwidths were initially estimated separately for each county using Scott’s rule (*h* = *σ* · *n*^−1/6^, where *σ* is the geometric mean of coordinate standard deviations and *n* is the number of spatial units). Scott’s rule yielded county-specific bandwidths of 10.4 km for Gaines County (n = 3 districts) and 4.4 km for Spartanburg County (n = 93 schools), reflecting the different spatial resolution of available data. Because county-specific bandwidths prevent direct comparison of density estimates (reviewer concern M2), we applied a common bandwidth of 0.066° (approximately 7 km), the geometric mean of the two county-specific estimates to both surfaces. This common bandwidth approximates a school catchment area radius and ensures that observed differences in peak density reflect genuine differences in susceptible clustering rather than bandwidth artifacts. Density surfaces were normalized to unvaccinated students per square mile and masked to county boundaries.

Bootstrap Uncertainty Quantification. To quantify uncertainty in peak susceptible density estimates, we implemented a parametric bootstrap procedure with n = 1,000 iterations. In each iteration, we perturbed the input data in three ways: (1) coverage perturbation: each school’s or district’s coverage rate was sampled from a normal distribution centered on the observed rate with standard deviation of ±3 percentage points for South Carolina (±5 percentage points for Texas, reflecting the additional uncertainty from cohort reconstruction); (2) spatial jitter: school coordinates were perturbed by a small random offset drawn from a bivariate normal distribution to account for geocoding uncertainty; and (3) enrollment sampling—the number of unvaccinated students was resampled from a binomial distribution given enrollment and perturbed coverage. For each bootstrap sample, we recomputed the KDE surface using the common 7-km bandwidth and recorded the peak susceptible density. The 2.5th and 97.5th percentiles of the bootstrap distribution define the 95% confidence interval. The ratio of Spartanburg-to-Gaines peak density was computed for each paired bootstrap sample to generate a distribution of density ratios with its own confidence interval. This approach propagates uncertainty from coverage measurement, geocoding, and enrollment stochasticity into the final density estimates.

#### Interpretation

The KDE visualization identifies spatial clustering of susceptible populations that may not be apparent from county-level summary statistics. In Spartanburg, multiple schools with moderate under-vaccination cluster together in the northwestern region of the county, creating dense pockets of susceptible students that can sustain transmission chains even when the county average appears acceptable. This spatial clustering helps explain why the South Carolina outbreak continues while the Texas outbreak was contained despite comparably sub-threshold coverage levels.

## Limitations

Several limitations should be considered when interpreting these data:

- Data structure asymmetry: Texas reports kindergarten-specific and 7th-grade MMR rates at the district level while South Carolina reports school-wide immunization rates across all grades at the school level. This introduces systematic differences in both the spatial resolution and age composition of coverage estimates between states. The comparison is between sub-county administrative units rather than equivalent school-level data.
- Cohort reconstruction assumptions: The Texas methodology assumes vaccination status at kindergarten entry and 7th grade remains stable through the corresponding grade span. Catch-up vaccination or delayed vaccination would lead to overestimation of current susceptibility, biasing the Texas unvaccinated count upward.
- Multi-grade school allocation: For 22 Spartanburg County schools spanning multiple grade levels, enrollment was allocated proportionally across levels by grade span. This assumes uniform immunization rates across all grades within a school, which may not hold if younger and older students have systematically different vaccination rates.
- Reporting unit differences: Texas data are reported at the district level (3 districts in Gaines County) while South Carolina data are reported at the school level (93 schools in Spartanburg County). The coarser Texas resolution means that within-district heterogeneity is unobserved.

